# The *IHME* vs Me: Modeling USA CoVID-19 Spread, Early Data to the *Fifth Wave*

**DOI:** 10.1101/2021.08.16.21262150

**Authors:** Genghmun Eng

## Abstract

Epidemiologists have never had such high-quality real-time pandemic data. Modeling CoVID-19 pandemic data became a predictive tool instead of an afterwards analysis. How early CoVID-19 model predictions impacted US Government policies and practices is first reviewed here as an important part of the pandemic history. It spurred independent modeling efforts, such as this, to help develop a better understanding of CoVID-19 spread, and to provide a substitute for the *IHME* (*Institute for Health Metrics & Evaluation, U. Washington*) 4-month predictions for the expected pandemic evolution, which they had to revise every couple of weeks. Our alternative model, which was developed over the course of several earlier *medrxiv.org* preprints, is shown here to provide a good description for the entire USA CoVID-19 pandemic to date, covering: (1) the original CoVID-19 wave [3/21/20-6/07/20], (2) the Summer 2020 Resurgence [6/07/20-9/25/20], (3) the large Winter 2020 Resurgence [9/25/20-3/19/21], (4) a small Spring 2021 “*Fourth Wave*”, [3/19/21-6/07/21], and (5) the present-day Summer 2021 “*Fifth Wave*” [6/07/21-present], which the USA is now in the midst of. Our analysis of the initial “*Fifth Wave*” data shows that this wave presently has the capacity to infect virtually all susceptible non-vaccinated persons who practice NO *Mask-Wearing* and minimal *Social Distancing*.

## 1 Introduction

The initial CoVID-19 pandemic response by the US Government and public agencies needs to be remembered for future pandemics. In spite of the availability of high-quality real-time pandemic data, the early history of the pandemic in the US shows that very little coordinated effort occurred to use this data in a timely manner, or to test the robustness of early CoVID-19 pandemic assumptions.

This report includes material from a recent presentation for the *Virtual MathFest 2021*, hosted by the *Mathematical Association of America (MAA)* on 3-7 August 2021. *Figure 1*, created by *Andy Balk* for the UK online news outlet *The Independent* ^**1**^, shows the US early CoVID-19 data combined with the US President’s early pandemic responses, including the President calling the pandemic a *“hoax”* that *“wasn’t out fault”*, before declaring a *National Emergency* in mid-March 2020.

**Fig. 1:**
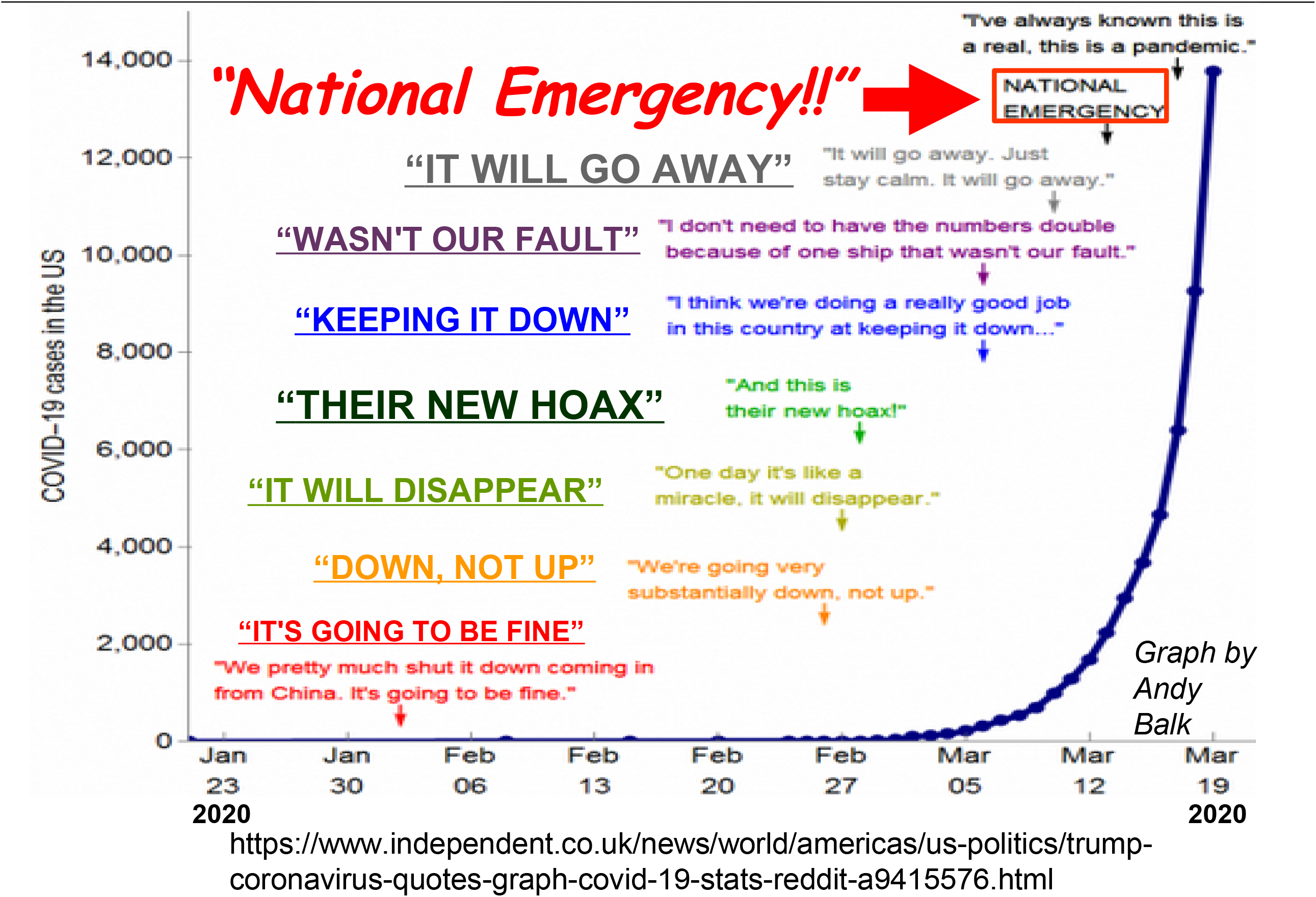
What President Trump Said in the Early Pandemic. Graph and text compiled by *Andy Balk* for the UK news outlet *The Independent*, showing the President minimizing the potential impacts of the pandemic, before reversing course and declaring it a *National Emergency* on 3/13/2020.

*Figure 2* captures the impact of early CoVID-19 pandemic modeling by the *University of Washington IHME (Institute for Health Metrics and Evaluation)*, which released a 4-month projection for CoVID-19 evolution in the USA^**2**^ on 3/25/2020. It was labelled *“America’s most influential coronavirus model”* ^**3**^, and it initially projected a total of about 81,000 CoVID-19 fatalities. Two weeks later, on 4/06/2020, the *IHME* revised its projections to only about 60,000 USA deaths by August 2020^**3**^. By 4/19/2020, the White House had adopted this *IHME* projection as a guide for their national policies^**4**^. Unfortunately, less than two weeks later, on 5/01/2020, the USA number of CoVID-19 deaths surpassed this value^**5**^, three months early. A day later, the internet blog *vox.com* offered the question^**6**^, *“The IHME coronavirus model keeps being wrong. Why are we still listening to it?”*

**Fig. 2:**
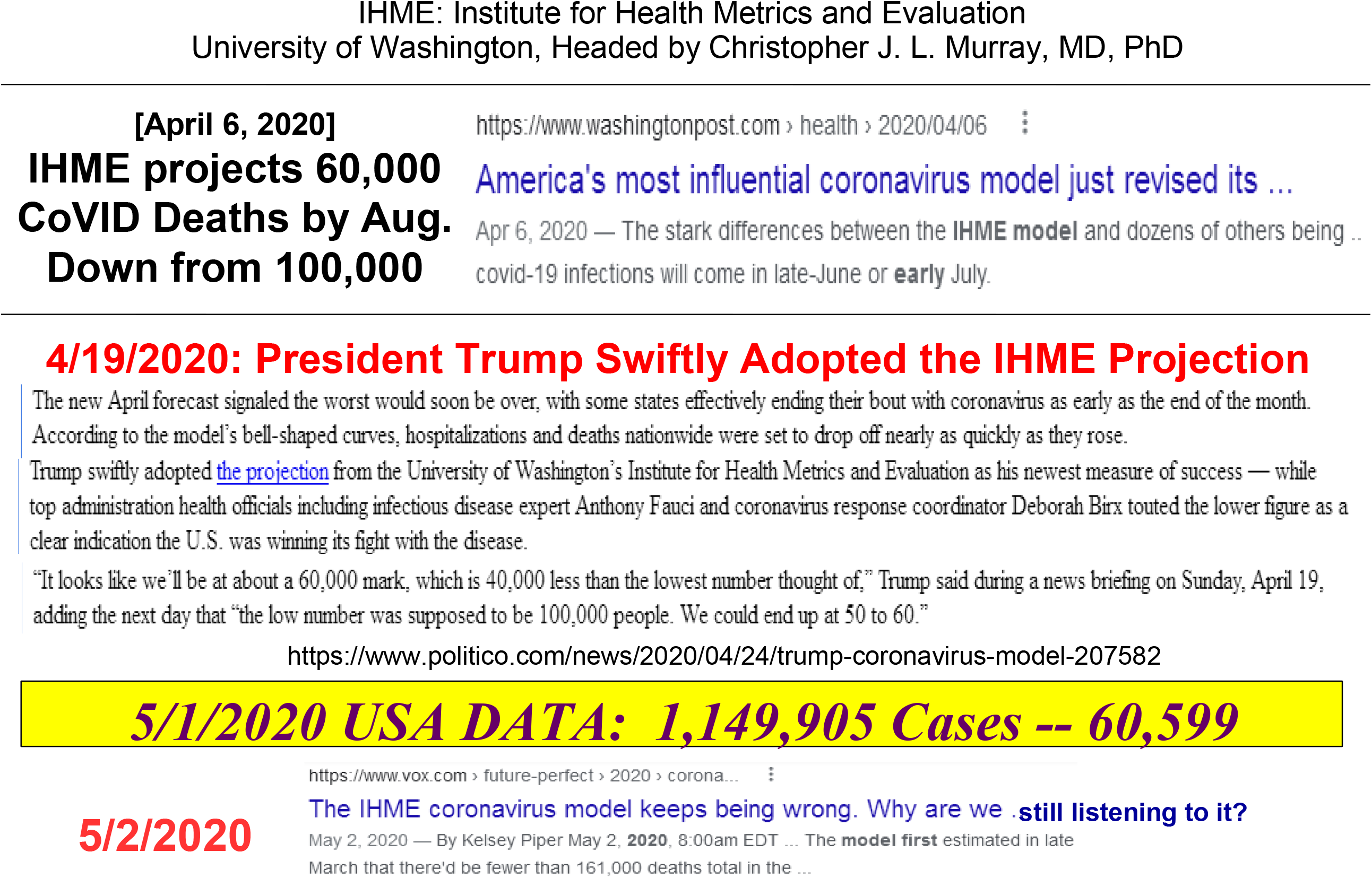
What the IHME Said in the CoVID-19 Early Pandemic. On 3/25/2020, the *University of Washington IHME (Institute for Health Metrics and Evaluation)* published their 4-month CoVID-19 projection, with two-week updates starting on 6 April 2020. The update projected only 60,000 USA CoVID-19 deaths by August 2020. These *IHME* projections quickly became “*America’s Most Influential Coronavirus Model* “. They were adopted by the White House on 4/19/2020 as part of the US Federal Government CoVID-19 response. Unfortunately, this 4-month future value was crossed on 5/1/2020, three months early, causing many to question the *IHME* modeling.

We found a persistent flaw in the *IHME* model. It is illustrated by the *IHME* graphs in *Figure 3*, which shows their *expected number of daily new cases* until CoVID-19 pandemic end, for both Northern Pennsylvania^**7**^ (*IHME*, 3/26/2020) and the nation^**8**^ (*IHME*, 4/22/2020). The persistent flaw is that the *IHME* **assumed** that symmetric functions for the *expected number of daily new cases* would always be applicable. They further **assumed** that these functions would be symmetric Gaussians, which drove all their predictions, as disclosed in their *MedRxiv* preprint of 3/25/2020^**9**^.

**Fig. 3:**
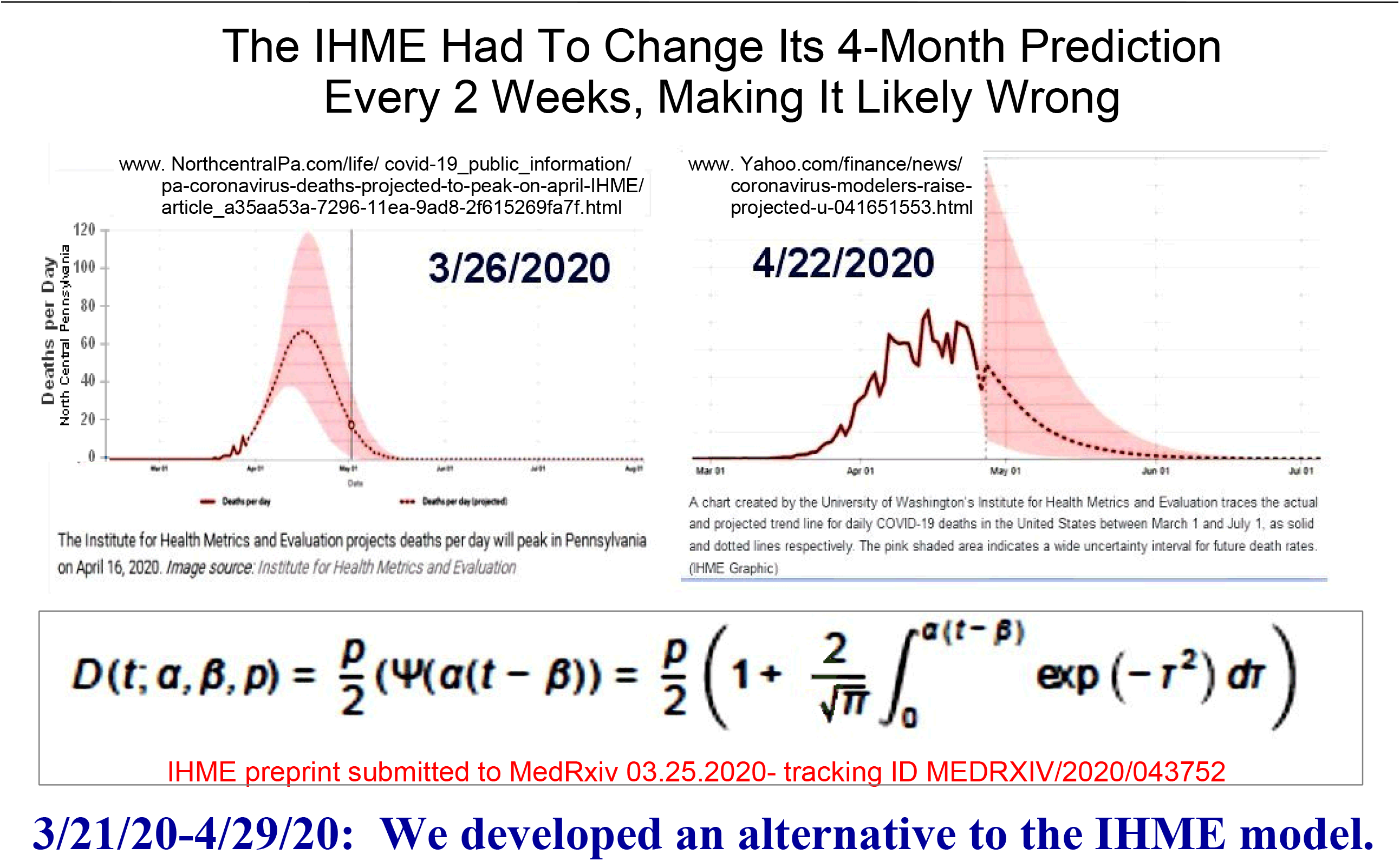
IHME Assumed Symmetric Functions Would Model the Rise and Fall of the Daily Number of Expected CoVID-19 Cases. *IHME* results for Northern Pennsylvania (3/26/2020) and the USA (4/22/2020), showing the *IHME* used the same symmetric function, pre-peak and post-peak, for the *expected number of daily new cases* [*dN* / *dt*] throughout this period. We found that this result arises solely due to the 3/26/2020 *IHME* model assumption that the *dN* / *dt* function had to be a Gaussian, which led us to develop a more data-based *IHME* model alternative.

Concerns about this *IHME* model and concerns with the Federal Government’s early CoVID-19 pandemic response resulted in many mathematically-minded individuals, including us, started their own independent CoVID-19 data analyses, while *Sewing Circles* all across America were making handmade masks for *First Responders* and patients.

The next section summarizes all of our CoVID-19 models, starting with our original March-April 2020^**10**^ *Social Distancing* model. This model was then expanded^**11-13**^ to include the likely effects of *Mask-Wearing*. A comparison of our model to the USA CoVID-19 pandemic data up through August 2021 shows that our expanded model is applicable to all USA CoVID-19 data, and provides a fairly accurate and quantifiable picture of the four USA CoVID-19 waves that have already occurred. Initial study of present-day “*Fifth Wave*” data also shows that it has the capacity to become the most deadly wave of all.

## 2 Initial Model Background and Development

This CoVID-19 study starts with the standard *SEIR (Susceptible, Exposed, Infected, Recovered/Removed)* model and *R*_0_-index, as shown in *Figure 4*. This *R*_0_ modifies exponentials so that *R*_0_ > 1 is pandemic growth; *R*_0_ < 1 is pandemic decay; while *R*_0_ = 1 gives a persistent disease baseline in the population.

**Fig. 4:**
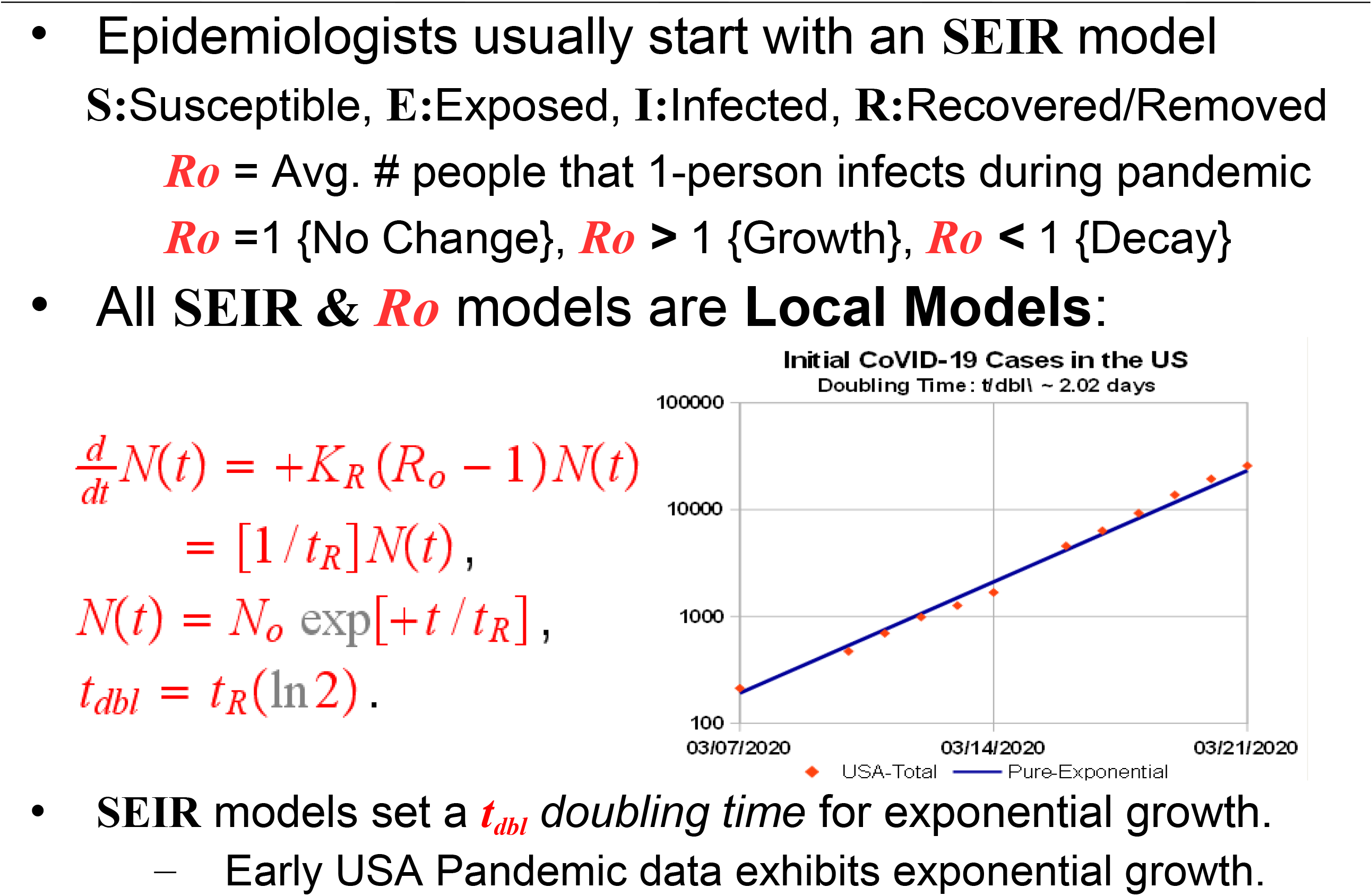
What *SEIR* Models and Pandemic *Ro* Factors Are. The standard *SEIR* Model (*Susceptible, Exposed, Infected, Recovered* or removed) is reviewed. It is a local model, which presumes an exponential growth for the total number of infected persons during the initial pandemic. Inset shows early CoVID-19 data matching this growth, giving a *doubling time* of *t*_*dbl*_ ≈ 2.02*d*.

Let *N*(*t*) be the *total number of pandemic cases*, with *dN* / *dt* being *the expected number of daily new cases*. The simplest *SEIR* model is:

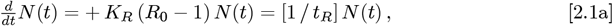

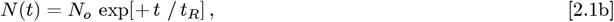

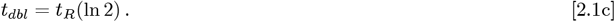

The {*K*_*R*_ *R*_0_} combination sets how fast an infected person spreads CoVID-19 to others; *N*_*o*_ is the number of infected people at time *t* = 0; and *t*_*dbl*_ is the pandemic *doubling time*. As shown in *Figure 4*, the early USA CoVID-19 data^**5**^ followed this exponential growth, with a very short *doubling time* of *t*_*dbl*_ ≈ 2.02 *d*. The form of the Eq. [2.1a] differential equation makes this *SEIR* model a *local model*, so it does not automatically account for changes in the collective behavior among a large population of *uninfected* people.

These collective behavior changes can arise from government mandates, or from new individual choices among *uninfected* people, such as *Social Distancing* and *Mask-Wearing*. These factors add a new **non-local** dimension to pandemic evolution, requiring extension of the Eqs. [2.1a]-[2.1c] *SEIR* exponential growth models.

On March 19, 2020, *Governor Gavin Newsom* of California ordered a CoVID-19 “stay-at-home” lockdown of virtually all of California’s ∼ 40 million residents^**14**^. Similar statewide CoVID-19 lockdowns were next ordered by the Governors of Illinois, New York, Indiana, Michigan, Ohio, Washington, West Virginia and Wisconsin^**15**^. Within days, this nationwide change slowed the CoVID-19 pandemic growth, as shown^**5**^ in *Figure 5*. This change corresponds to an increasing *t*_*R*_ or *t*_*dbl*_, making these parameters time-dependent. We next showed^**10**^ that the simplest *t*_*dbl*_(*t*) model, using a linear function for *t*_*dbl*_(*t*):

**Fig. 5:**
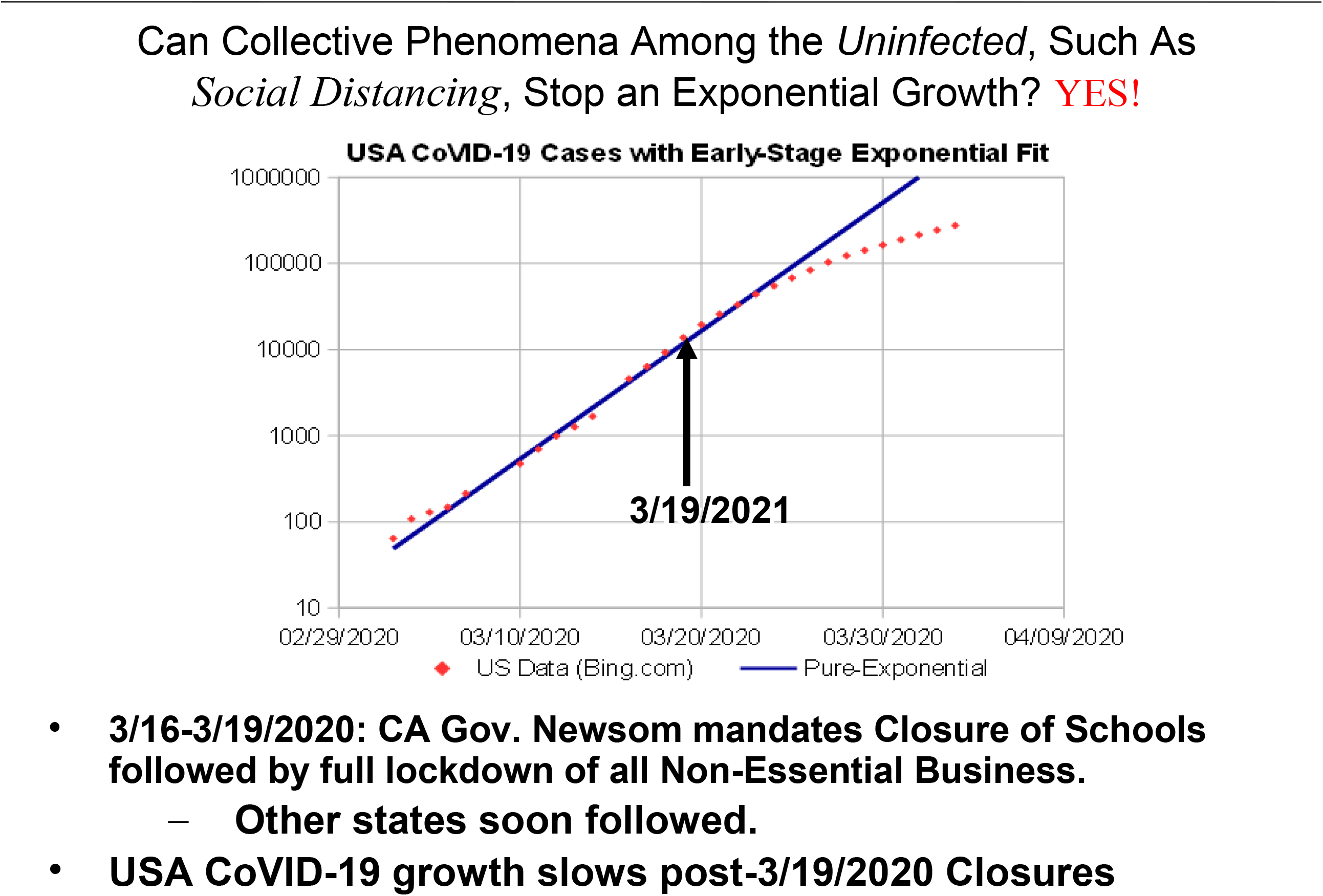
*Social Mitigation Measures* Slow Down CoVID-19 Growth. Within days of starting wide-scale *Social Distancing* measures among *uninfected* persons, including closure of schools and lockdown of non-essential businesses, the pandemic *t*_*dbl*_ *doubling time* lengthened and CoVID-19 growth slowed.

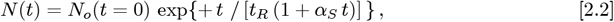

naturally predicts pandemic end, since:

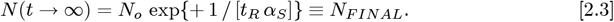

The *α*_*S*_ parameter measures the amount of *Social Distancing*. Instead of being a symmetric function, the *dN* / *dt* from Eq. [2.2] for *the expected number of daily new cases* is very asymmetric, with a predicted ^∼^1/*t*^2^ long-term tail:

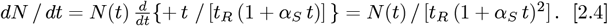

This asymmetry in *dN* / *dt* closely matches the *bing.com* CoVID-19 USA data up through 4/19/2020^**17**^, as shown in *Figure 6*.

**Fig. 6:**
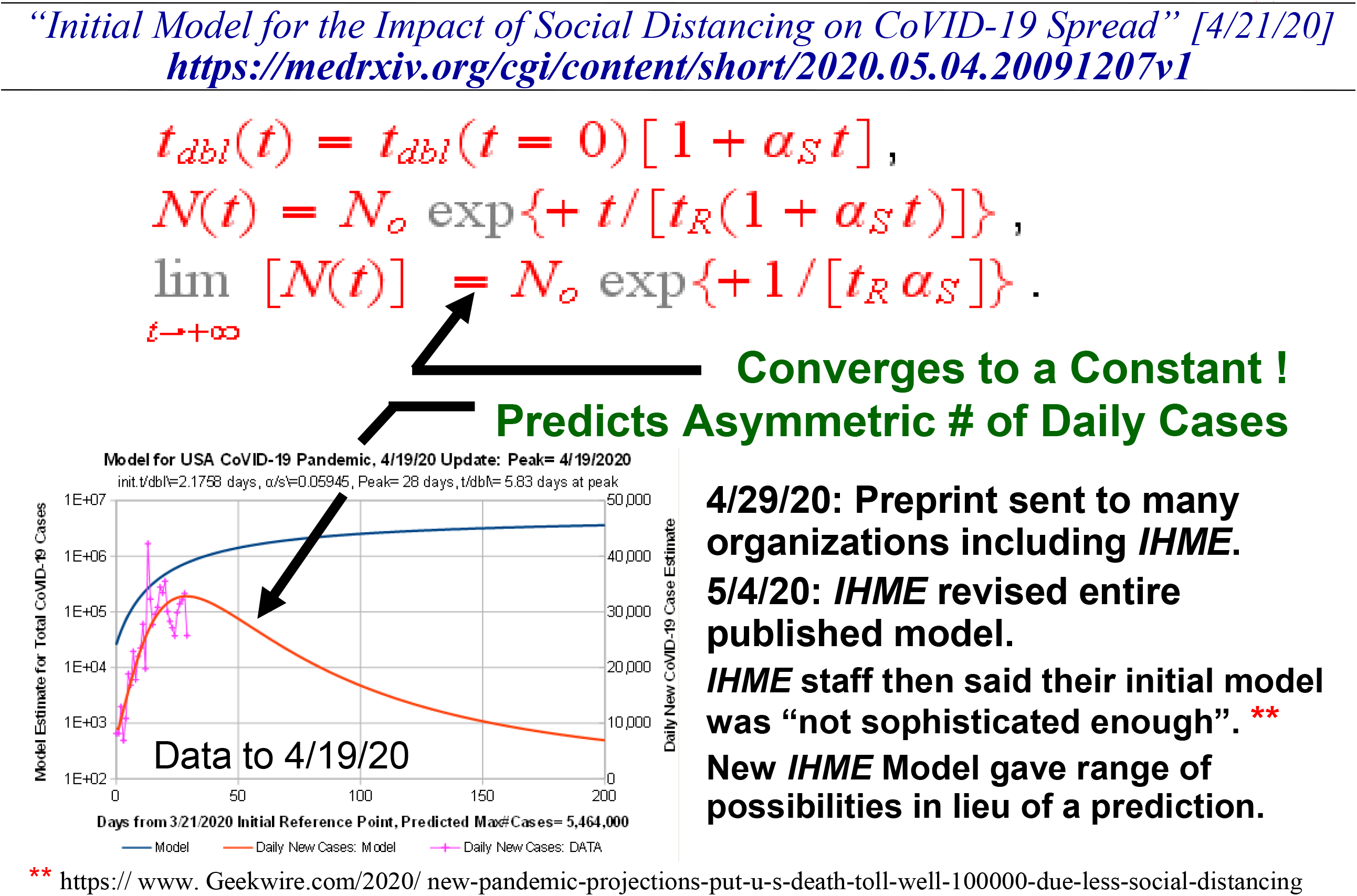
The Simplest CoVID-19 Model: Let *t*_*dbl*_ → *t*_*dbl*_(*t*). Collective phenomena among the *uninfected*, such as *Social Distancing*, can stop an exponential growth if *t*_*dbl*_(*t*) itself is a linear function of time, so that *N*(*t* → ∞) automatically converges to a finite value. A data vs model comparison to 4/19/2020 shows that a highly asymmetric *dN* / *dt* is predicted. On 4/29/2020, our *MedRxiv* pre-print covering this original model development and data analysis was sent to many organizations, including the *IHME*. Within a week, the *IHME* staff substantially revised their entire reported modeling methods, saying that their original CoVID-19 model was not “*sophisticated enough*”.

Our original preprint was sent to multiple organizations on 4/29/2020, including the *IHME*. Within a week, on 5/4/2020, the *IHME* substantially revised their entire reported modeling effort^**16**^, and offered only a range of possibilities, in lieu of a specific prediction. As reported by *Alan Boyle* of *geekwire.com*^**17**^, the *IHME* researchers acknowledged on 5/4/2020 that their previous CoVID-19 modeling wasn’t “*sophisticated enough*”.

*Figure 7* shows the results of applying Eq. [2.2] to the ongoing *bing.com* CoVID-19 USA data up through 6/07/2020, seven weeks later than in *Figure 6*. The {*N*_*o*_[3/21/2020] *t*_*R*_, *α*_*S*_} values changed only by 8% –10% between the 4/19/20 and 6/07/20 analyses. The *N*_*FINAL*_ value for the total number of USA CoVID-19 cases predicted at pandemic end, changed ^∼^18%, from *N*_*FINAL*_ ≈ 5.46 *million* to *N*_*FINAL*_ ≈ 4.50 *million*. It represents a significantly smaller model prediction change over a much longer time, compared to the 3/25/2020 *IHME* model.

**Fig. 7:**
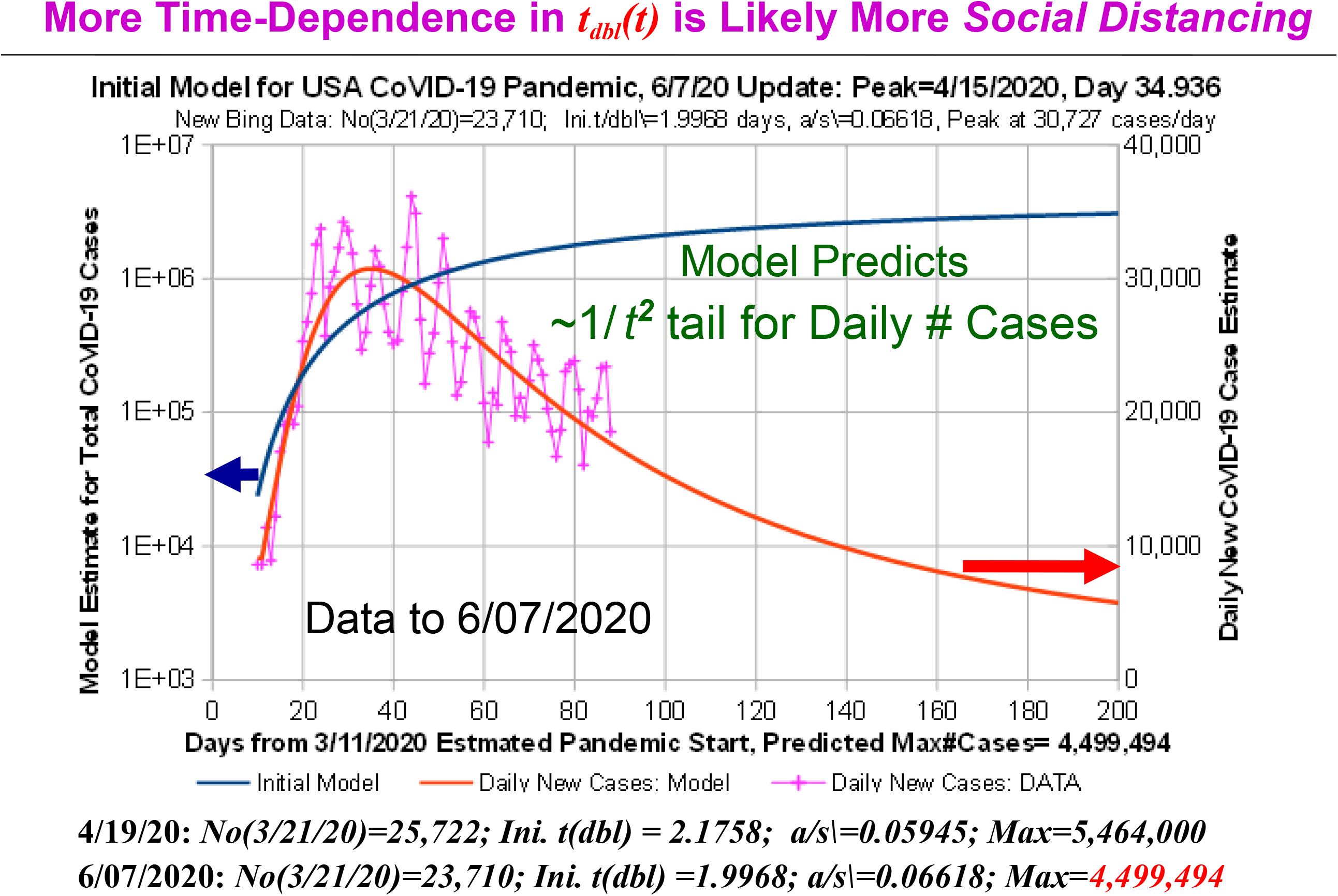
Same Model Gives Similar Predictions with Later USA Data. Our model was applied to the later USA CoVID-19 pandemic data, up through 6/07/2020, nearly 7 weeks after *Figure 6*. The parameter values changed only about 10% or less between these two periods, which altered the USA *N*(*t* → ∞) value about 18%, from ∼5.464 to ∼4.499 *million*.

*Figure 8* shows the world-wide early CoVID-19 data, as assembled by *The Royal Society of London* for their early CoVID-19 pandemic 8/24/2020 review^**18**^. Various preset *t*_*dbl*_ values are also shown as a visual guide. Using a logarithmic ordinate, virtually all these data show a downward curvature away from the straight-line function of pure exponential growth.

**Fig. 8:**
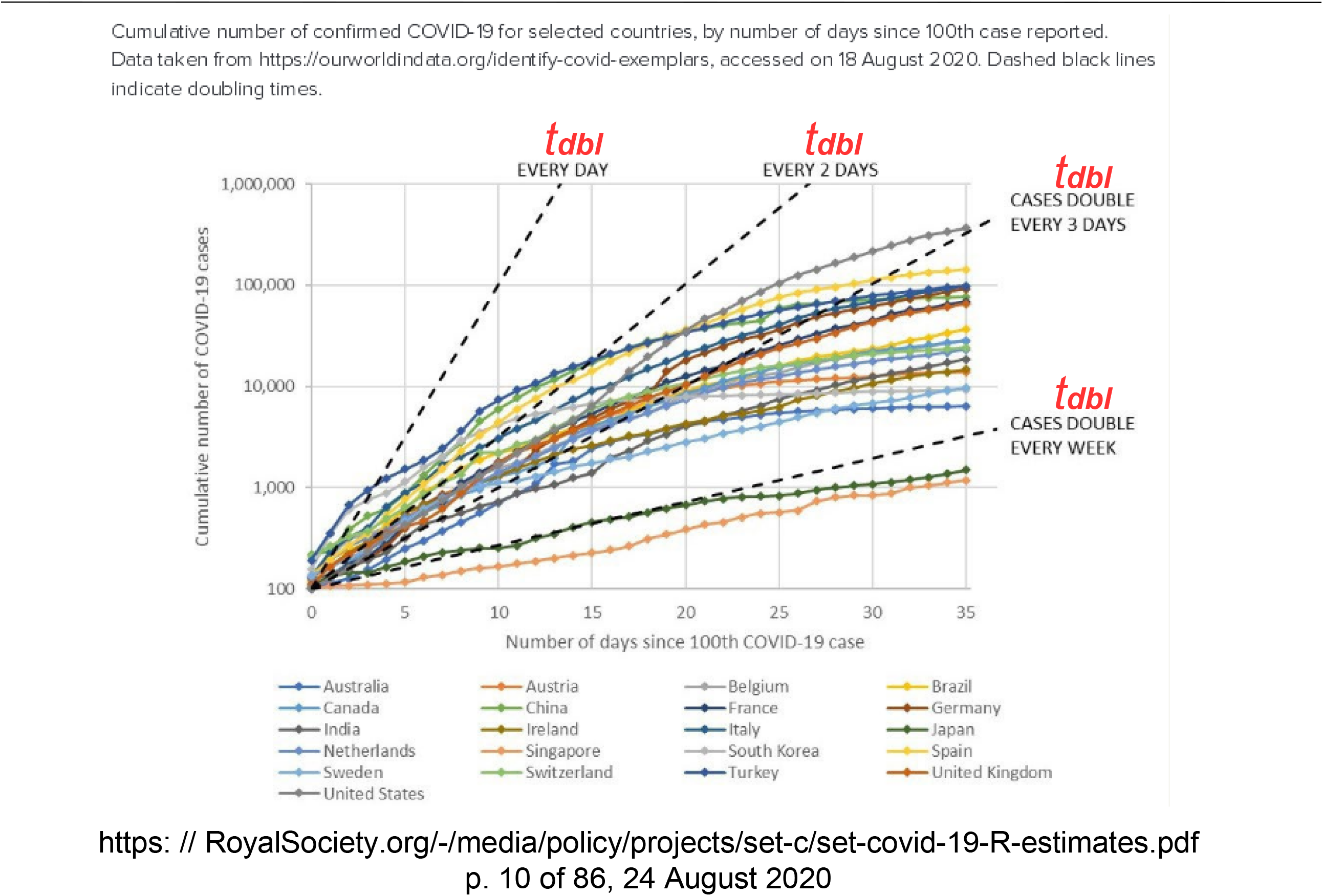
Plot of World-Wide CoVID-19 Early Pandemic DATA. Graph by the *Royal Society of London* from their 8/24/2020 early CoVID-19 pandemic review. On a logarithmic ordinate, virtually all these data show a downward curvature from the straight-line of pure exponential growth.

In *Figure 9*, the Eq. [2.2] model was applied to the early pandemic growth for various countries and the World, showing it provided a fairly good approximation for most cases examined^**10**^. However, as *Figure 10* shows, the early CoVID-19 data from Italy was somewhat different, in that it had a post-peak *dN* / *dt* tail that was almost a pure exponential decay down to fairly low values, which made it difficult for the Eq. [2.2] model to handle.

**Fig. 9:**
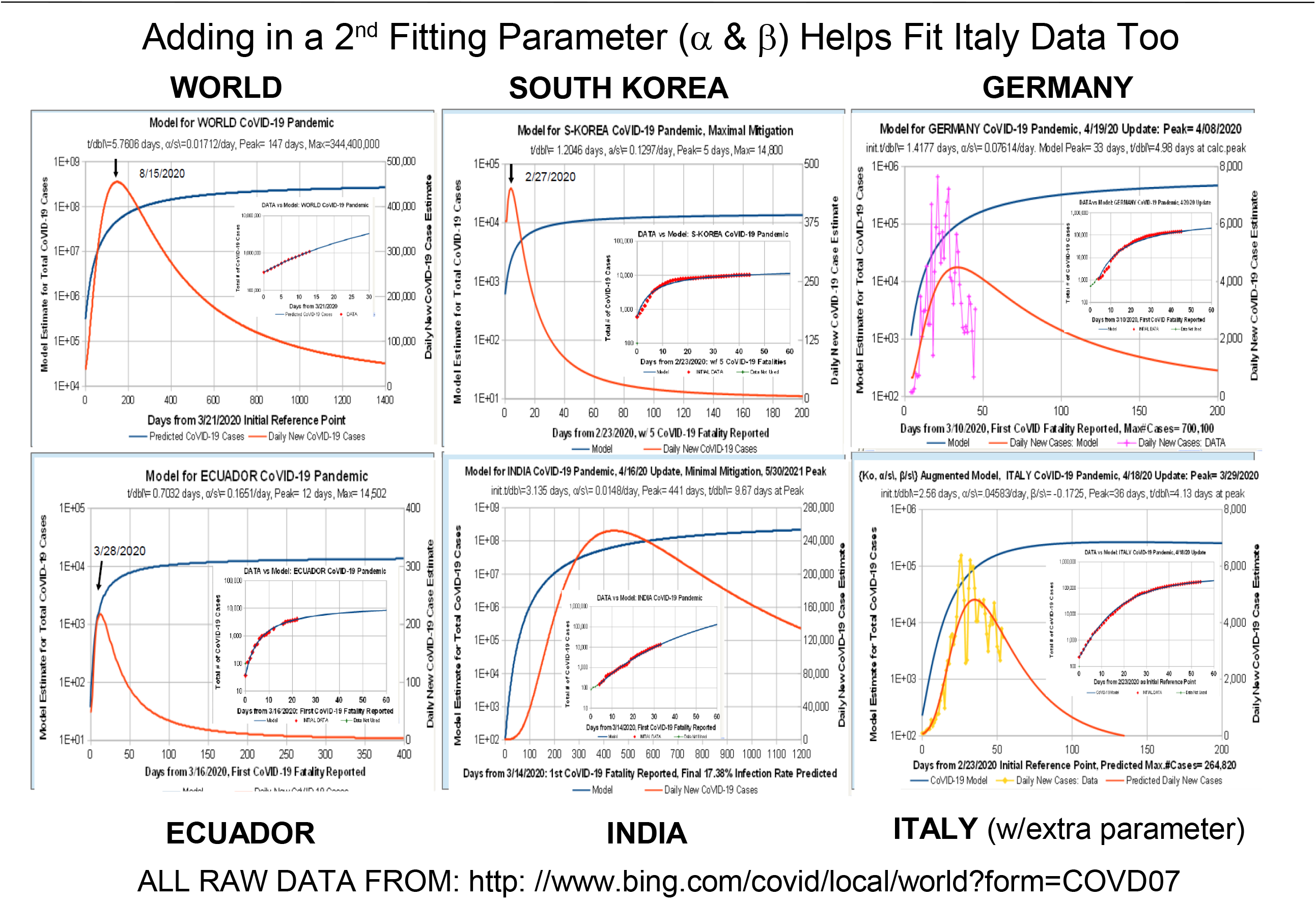
Model Approximates Early Pandemic World Data Except for Italy. Early pandemic data up through 4/19/2020, with model datafits are shown for the World and various countries. Our model worked fairly well on early CoVID-19 pandemic data for many countries except for Italy.

**Fig. 10:**
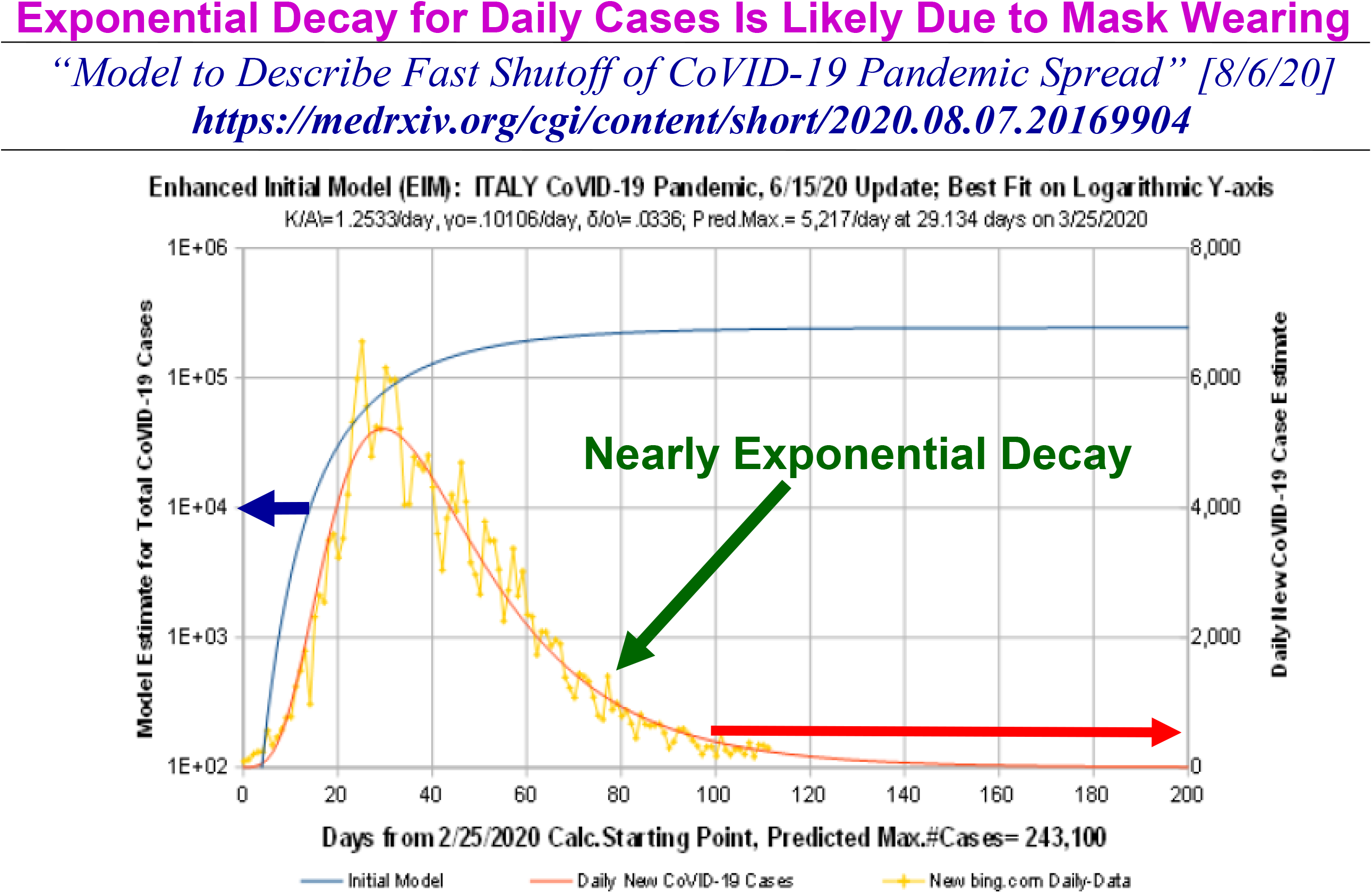
Updated Model for Italy. Empirically, the initial wave post-peak *dN* / *dt* data for Italy decreased nearly exponentially, which is outside the realm of *dN* / *dt* ∼ 1 */ t*^2^ function, associated with our which original CoVID-19 pandemic model. As reported by NPR, Italy achieved this result with help from the *People’s Republic of China*, which recommended public *Mask-Wearing*, making Italy the first country where this was mandated.

As *NPR* reported^**19**^, the *People’s Republic of China*, as part of their early CoVID-19 pandemic assistance to Italy, recommended three large-scale changes, which the Italian government quickly adopted: (1) significant mask-wearing, (2) more aggressive business shutdowns, and (3) more social distancing. It is doubtless that these aggressive changes contributed to a different early CoVID-19 pandemic evolution in Italy.

A second parameter was then added to the Eq. [2.2] model^**12**^, to explicitly enable *dN* / *dt* long-term tails to exhibit a nearly exponential decay with time:

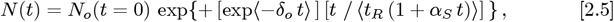

but it needs an added restriction that this pandemic wave ends whenever the calculated *dN* / *dt* < 0 first occurs using Eq. [2.5].

Using Eq. [2.5] gives the Italy datafit shown in *Figure 10*. Since the main difference between the USA and Italy CoVID-19 responses at that time was significant *Mask-Wearing*, the *δ*_*o*_ size is likely to primarily be a *Mask-Wearing* metric.

Follow-on analyses for the successive waves of USA CoVID-19 infections are shown in *Figures 11-15*. In addition to covering: (a) the initial Spring 2020 pandemic [*Figure 7*], this Eq. [2.5] few-parameter model reasonably fits: (b) the Summer 2020 Resurgence [*Figure 11*], (c) the large Winter 2020 Resurgence [*Figures 12* and *13*], (d) the smaller Spring 2021 “*Fourth Wave*” [small *dN* / *dt* peak in *Figures 12* and *13*], and (e) the present-day Summer 2021 “*Fifth Wave*” [*Figures 14-15*].

**Fig. 11:**
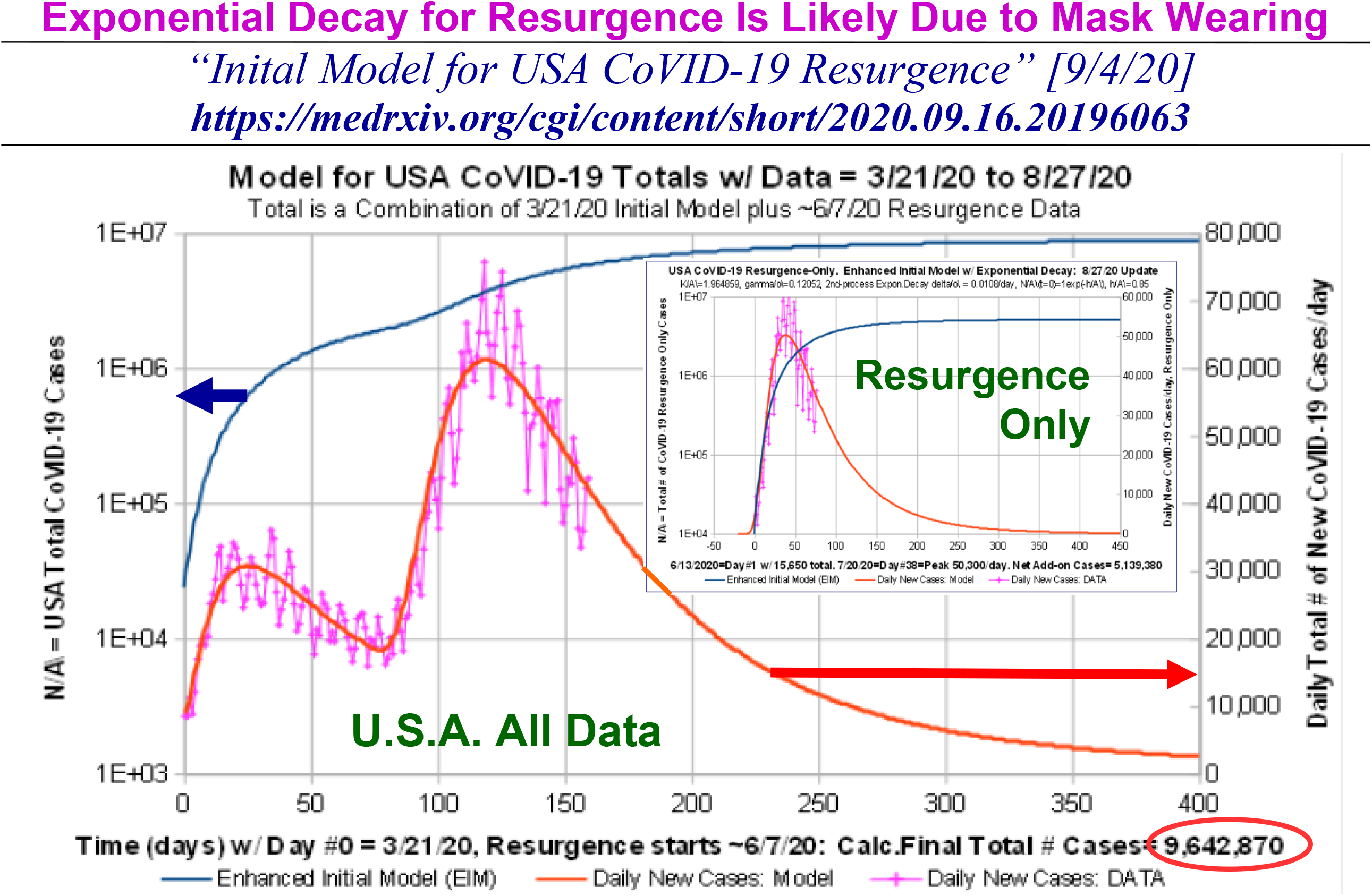
Modeling the Summer 2020 USA Resurgence. The Eq. [2.5] extended model has two parameters {*α*_*S*_; *δ*_*o*_} for quantifying the collective behavior of *uninfected* persons in enabling CoVID-19 pandemic shutoff. infections. One is associated with *Social Distancing* and the other is associated with *Mask-Wearing*. The USA Summer 2020 CoVID-19 Resurgence by itself (inset) shows that the *dN* / *dt* post-peak tail has an exponential decay component, similar to the prior Italy data. That period also corresponds to the first time there was significant USA Mask-Wearing. The USA *N*(*t* → ∞) pandemic-end value increased from ∼4.499 *million* (initial wave) to ∼9.643 *million* (initial wave plus Summer 2020 Resurgence).

**Fig. 12:**
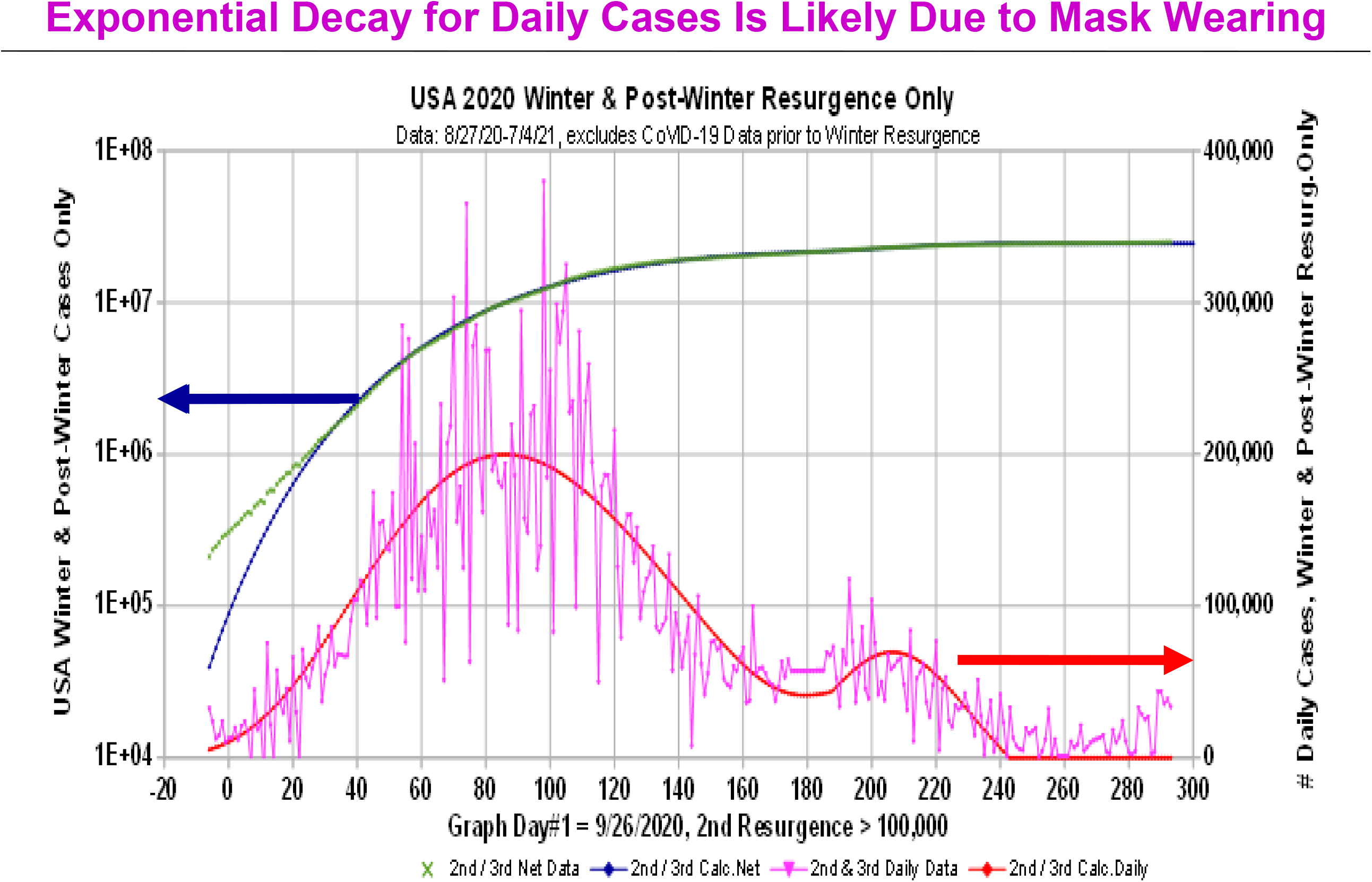
Modeling the Large Winter USA 2020 Resurgence By Itself. This Figure shows the long USA Winter 2020 Resurgence by itself, after subtracting out both the Spring 2020 initial wave and Summer 2020 Resurgences. Because of the size and duration of this Winter Resurgence, it spilled over into early 2021. A smaller Spring 2021 “*Fourth Wave*” is also evident.

**Fig. 13:**
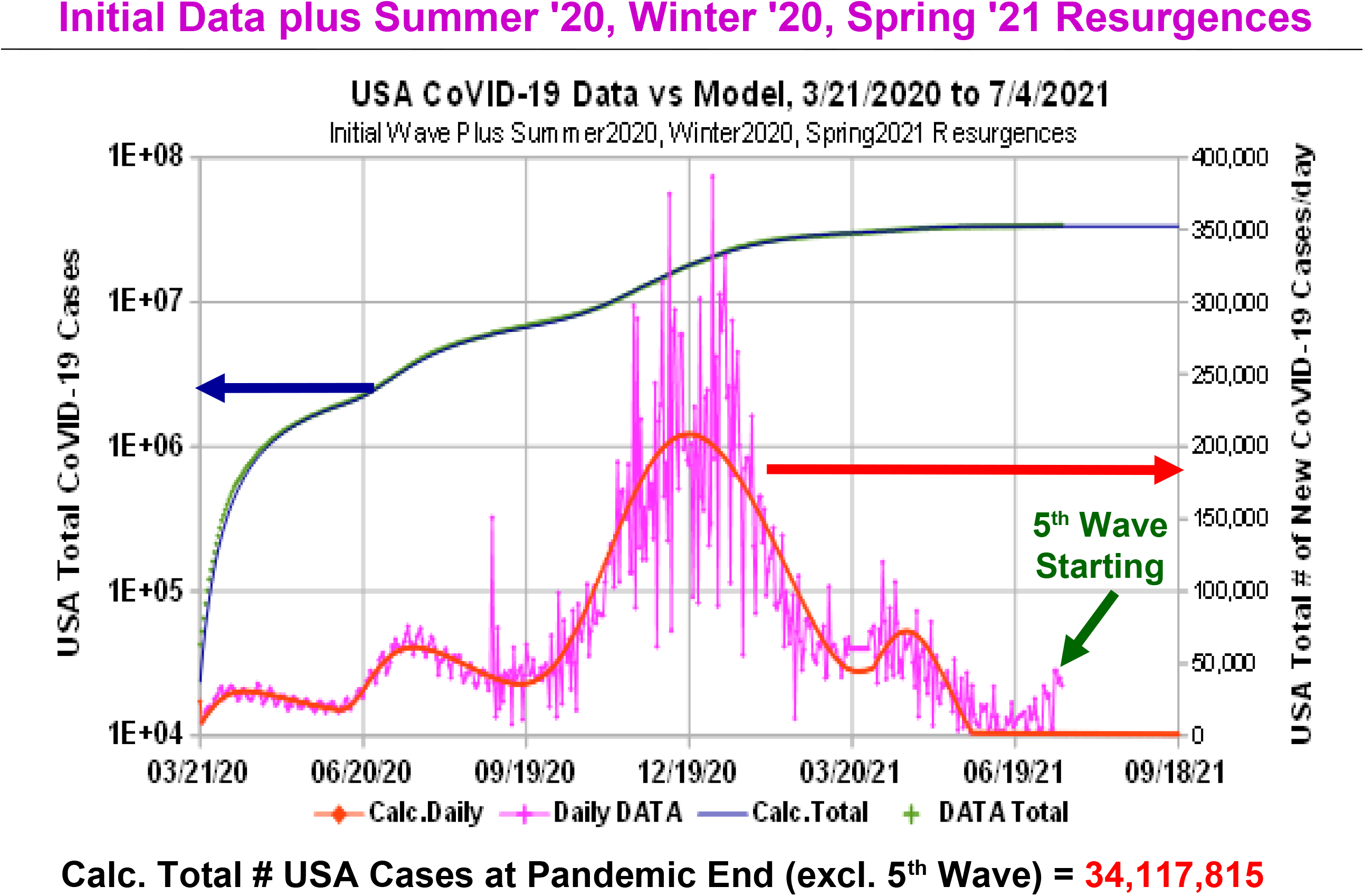
Modeling 2020-2021 USA Totals for CoVID-19 Pandemic. The total number of USA CoVID-19 infections [*N*(*t*), left axis] and the *expected number of daily new cases* [*dN* / *dt*, right axis] are shown, from the 3/2020 start of wide-scale *Social Distancing* to the present-day 8/2021, although model fits only used data up to 7/4/2021. The rise in the CoVID-19 *daily new cases* after that signals a new Summer 2021 “*Fifth Wave*”. Each CoVID-19 wave is assumed to end at the first calculated *dN* / *dt* < 0 point for that wave, which also provides a good marker for the next CoVID-19 start. USA infections at pandemic-end [*N*(*t* → ∞)] now exceeds 34 *million*.

**Fig. 14:**
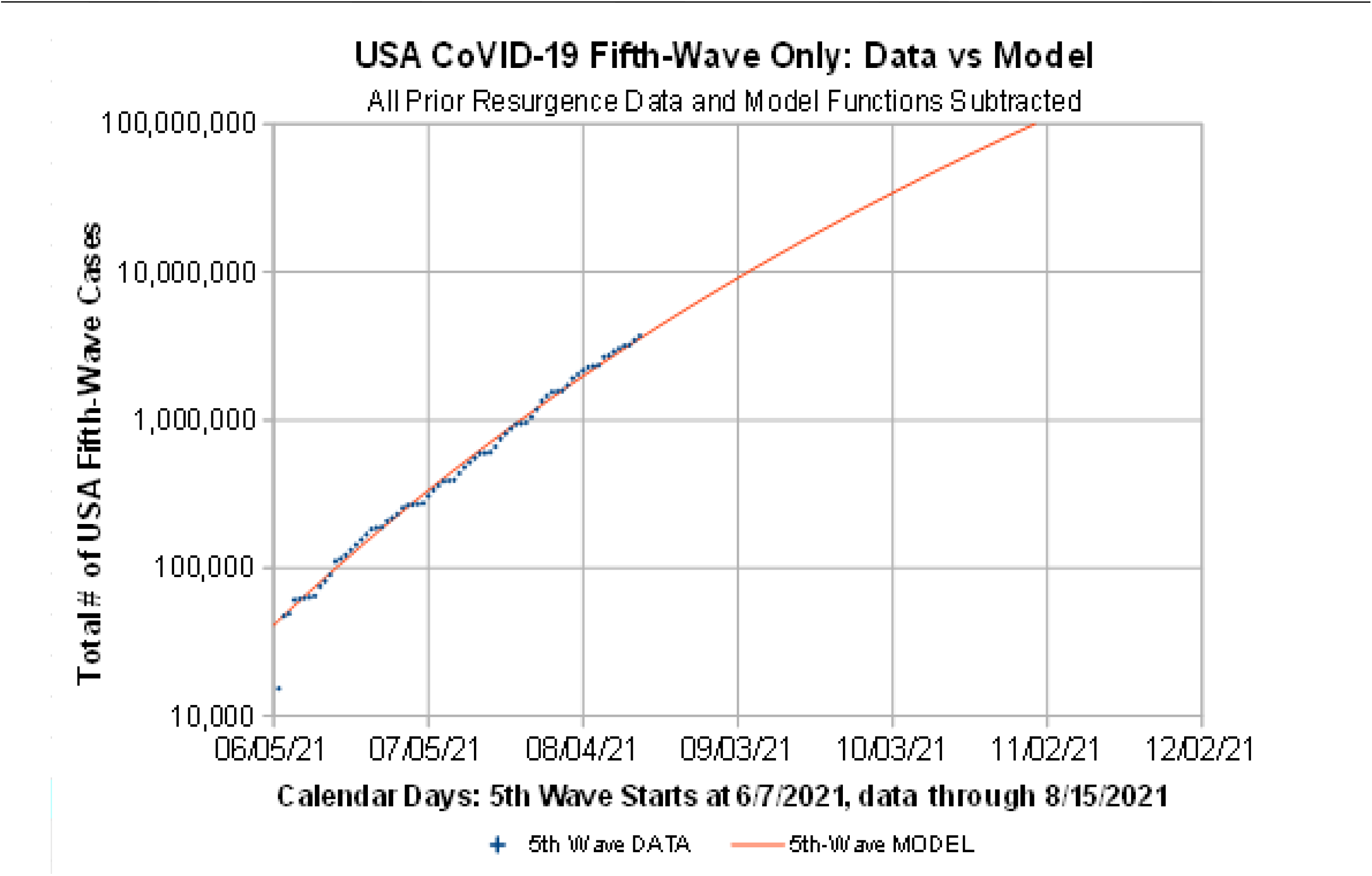
Initial Analysis of USA CoVID-19 “*Fifth Wave*” By Itself. This graph plots the *N*(*t*) number of “*Fifth Wave*” cases by itself, with all prior CoVID-19 waves removed. Our estimated start date for this “*Fifth Wave*” is 6/07/2021. The relative lack of downward curvature, as compared to the *Figure 8* data, shows that this “*Fifth Wave*” has very little *Social Distancing* or *Mask-Wearing* among this newly infected sub-group.

Since each new CoVID-19 wave sits atop the tails from all the prior CoVID-19 waves, the CoVID-19 overall progression in *Figure 15* shows that this multi-wave analysis has retained its overall validity, from the initial 3/21/2020 start date, up through the latest data. Model parameters that were derived from each prior wave did not need any revisions, once the next new CoVID-19 wave became established as exceeding the prior baseline.

**Fig. 15:**
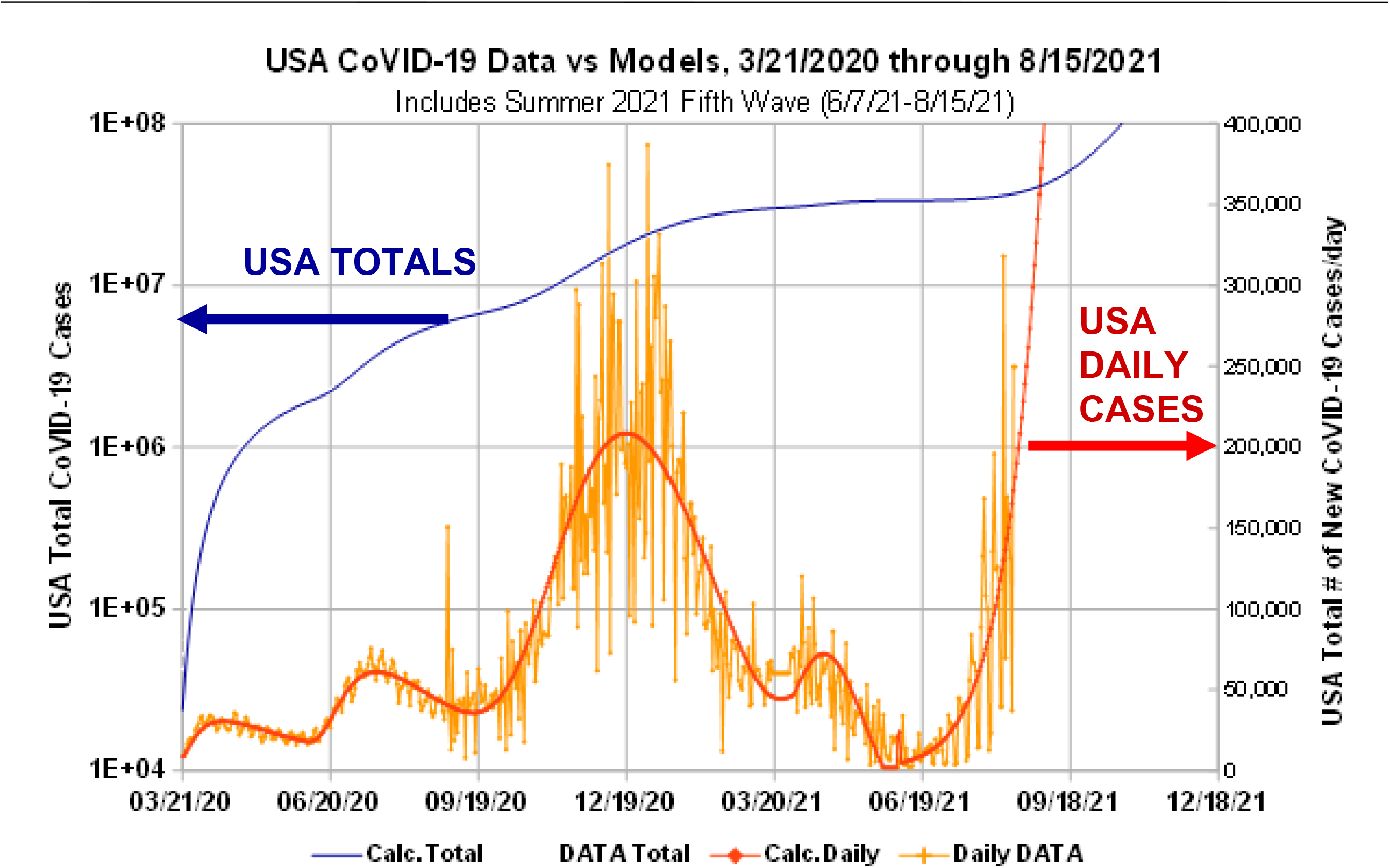
USA CoVID-19 Totals Including Early “*Fifth Wave*” Data. The blue curve shows the *N*(*t*) projections for all the USA CoVID-19 waves, including this “*Fifth Wave*”. The daily *dN* / *dt* values are in red, and are projected to continue to rise sharply.

The various {*t*_*R*_, *α*_*S*_, *δ*_*o*_}values associated with each of these CoVID-19 stages are summarized in *Figure 16*. Those parameter values provide additional insight as to what factors were likely driving the CoVID-19 infection rate for each USA pandemic wave.

**Fig. 16:**
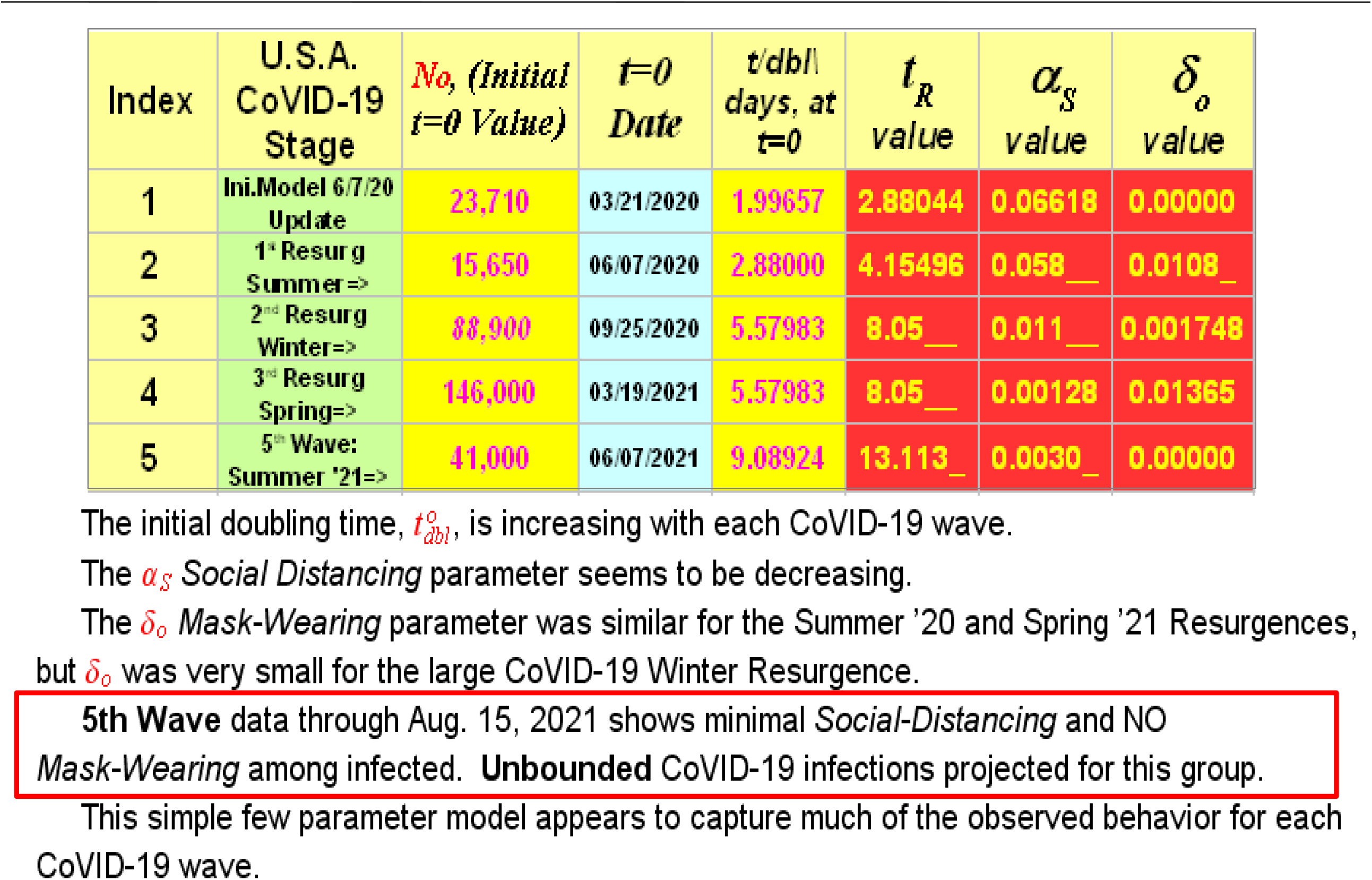
Summary of CoVID-19 Models and Model Parameters. This Table summarizes model parameters for each USA CoVID-19 pandemic wave. The *t*_*R*_ or *t*_*dbl*_(*t* = 0) *doubling times* at the start of each CoVID-19 wave are increasing, but larger *t*_*R*_ values only delay the time when all susceptible people become infected. More *Social Distancing* or *Mask-Wearing* is needed to prevent the present-day USA “*Fifth Wave*” from infecting all susceptible persons. See text *Section 3* of the main text for additional interpretation of these results.

## 3 What the Parameter Values Indicate

The first weeks of the CoVID-19 pandemic (3/7/2020-3/21/2020) had an *N*(*t*) *doubling time* of ∼ 2.02 *d*, as shown in *Figure 4*. As given in the *Figure 16* table, the entire pandemic first-wave, from 3/21/2020 through 6/07/2020, showed a best-fit value of *t*_*R*_ = 2.8804, corresponding to an intrinsic *doubling time* of:

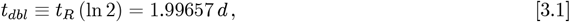

which is virtually identical to the original *Figure 4* early-pandemic value. Since this *t*_*R*_ parameter is closest to the Eqs. [2.1a]-[2.1c] basic *SEIR* model, it shows that the intrinsic USA-wide CoVID-19 spreadability remained constant. Thus, virtually the entire USA pandemic mitigation for this time interval was due to the *Social Distancing* measures that were implemented. The calculated value of *α*_*S*_ = 0.066l8 */ day* gives a final CoVID-19 pandemic end of *N*_*FINAL*_ ≈ 4.4995 *million* USA cases total.

Unfortunately, loosening controls led to a CoVID-19 Summer 2020 Resurgence, which likely started around 6/07/2020, 78 days after the start of USA-wide *Social Distancing* on 3/21/2020. Other factors such as different CoVID-19 variants circulating could have also contributed to the overall size of this resurgence. The Eq. [3.1] *t*_*R*_-value and intrinsic *doubling time* for the Summer 2020 Resurgence was 40% – 50% larger than the CoVID-19 initial wave, indicating that *SEIR* parameters for this resurgence were actually less aggressive. The calculated *Social Distancing* value of *α*_*S*_ = 0.058 / *day* was similar to the initial CoVID-19 pandemic wave, indicating a similar amount of *Social Distancing* mitigation.

People in the USA started to engage in *Mask-Wearing*, so that the post-peak decline for this Summer 2020 resurgence was faster than for the initial Spring 2020 CoVID-19 wave, as shown by the *Figure 11* inset. Our analysis gave *δ*_*o*_ = 0.0108 / *day* for *Mask-Wearing*. Although this value is significantly smaller in size than *α*_*S*_, its unit-for-unit impact is greater. Thus, *Mask-Wearing* is more powerful as a CoVID-19 pandemic stopping agent than *Social Distancing*. Had all of those controls remained in place, and had the weather not changed, these calculations would have predicted a CoVID-19 pandemic end with *N*_*FINAL*_ ≈ 9.643 *million* USA cases total.

But the seasons changed, the USA weather changed, and the dreaded, almost unavoidable, Winter 2020 Resurgence then occurred. Our best estimate for its starting date was 9/25/2020, 110 days after the 6/07/2020 start of the Summer 2020 Resurgence. Its Eq. [3.1] *t*_*R*_-value and intrinsic *doubling time* was almost 2*X* larger than the Summer 2020 Resurgence, indicating that *SEIR* parameters continued to become even less aggressive. However, both the *Social Distancing* parameter *α*_*S*_, and the *Mask-Wearing* index *δ*_*o*_ were much less, which made the Winter 2020 Resurgence very long and very deadly.

It overlaps with a small Spring 2021 “*Fourth Wave*”, which started around 3/19/2021, 175 days after the Winter 2020 Resurgence started on 9/25/2020. This small “*Fourth Wave*” likely ended 0 days later, around 6/07/2021, which ironically covered virtually the same calendar interval as the original CoVID-19 pandemic start. Combining the Winter 2020 Resurgence and this small “*Fourth Wave*”, with the prior CoVID-19 waves now predicted a final CoVID-19 pandemic end at *N*_*FINAL*_ ≈ 34.8l7 *million* USA cases total.

We are now in the CoVID-19 pandemic “*Fifth Wave*”. We estimate that it started around 6/07/2021, and it is a pernicious Summer 2021 Resurgence. Our initial analyses shows that the basic *SEIR* model *t*_*R*_ parameter continues to increase, which is good. But any positive *t*_*R*_ value means that the CoVID-19 pandemic can still exponentially grow at any time. CoVID-19 pandemic shutoff needs a relatively large *α*_*S*_ *Social Distancing* value, and a finite *Mask-Wearing* index *δ*_*o*_ to hasten the pandemic end.

Our model tells us who is getting infected in this “*Fifth Wave*”. As the *Figure 16* Table shows, the infected people in this “*Fifth Wave*” are associated with virtually NO *Mask-Wearing*, and very little *α*_*S*_ *Social Distancing*. The *α*_*S*_ and *δ*_*o*_ values are so small for the beginning of this “*Fifth Wave*” that our model predicts an *N*_*FINAL*_ value in the *billions*, assuming an arbitrarily large susceptible population. What such a large *N*_*FINAL*_ means is that, unabated, this “*Fifth Wave*” can infect virtually every susceptible non-vaccinated person in USA who is in this “minimal *Social Distancing*” and “NO Mask-Wearing” sub-group.

## 4 Summary

Let *N*(*t*) be the *total number of pandemic cases*, with *dN* / *dt* for *the expected number of daily new cases. Figures 1-3* here summarize elements of the early USA CoVID-19 response. One of the most influential early USA CoVID-19 models by the *IHME (Institute for Health Metrics and Evaluation)* was likely wrong, which led us to develop an alternative model for USA CoVID-19 spread.

For the early CoVID-19 pandemic, a basic *SEIR (Susceptible, Exposed, Infected, Recovered/Removed)* epidemiology model accurately predicted an *N*(*t*) exponential growth :

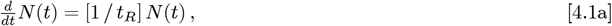

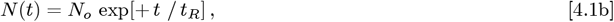

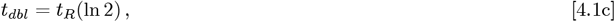

where *t*_*R*_ is the pandemic growth rate, and *t*_*dbl*_ is the *N*(*t*) *doubling time*.

Because *SEIR* models are *local models* that track and predict the number of infected persons, they do not automatically account for changes in the collective behavior among the large population of *uninfected* people. These changes can be due to new government mandates, or changes in the individual choices among *uninfected* people, such as *Social Distancing* and *Mask-Wearing*. These factors can add a new non-local dimension to pandemic evolution, requiring an extension of the basic Eqs. [4.1a]-[4.1c] *SEIR* exponential growth models.

The USA CoVID-19 data showed that these large-scale interventions convert *t*_*dbl*_ in Eq. [4.1c] into a function of time *t*_*dbl*_(*t*). To model these changes, early data supported using a nearly linear function of time for *t*_*dbl*_(*t*):

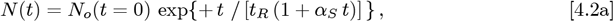

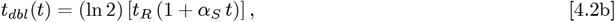

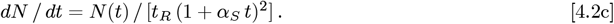

A simple interpretation of *α*_*S*_ is that it measures the amount of *Social Distancing* among *uninfected* persons, as a pandemic mitigation measure. When *t*_*dbl*_(*t*) is a nearly linear function of time, the total number of infected persons at the pandemic end, *N*(*t* → ∞), will converge to a finite value, automatically shutting the pandemic off. Having a sub-linear *t*_*dbl*_(*t*) growth would only slow the pandemic down, but not stop it from eventually infecting all susceptible persons. *Social Distancing* is needed, and it has to be robust enough so that the long-time behavior of *t*_*dbl*_(*t*) is at least a linear function of time to enable pandemic shutoff.

This model provided a good fit to the early USA CoVID-19 data, holding its predictive power for nearly two months. The Eqs. [4.2a]-[4.2c] functions were found to provide acceptable CoVID-19 datafits for many other countries around the world, except for Italy.

Italy differed from many other countries that mandated *Social Distancing, Non-Essential Business Closures*, and *Lockdowns*, in that they also instituted significant *Mask-Wearing*. As a result, their post-peak decline in *dN* / *dt* for their first major CoVID-19 wave showed nearly an exponential decay, which is significantly faster than the Eq. [4.2c] predictions. A second parameter was added to Eq. [4.2a], giving:

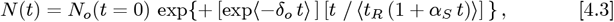

to cover this new case, but an additional restriction is needed that the end of the pandemic wave occurs when *dN* / *dt* < 0 is first calculated in Eq. [4.3]. The simplest interpretation for this *δ*_*o*_ second mitigation parameter is that it measures the amount of *Mask-Wearing* among *uninfected* persons.

While the Eq. [4.2a] model fits (a) the initial USA Spring 2020 pandemic [*Figure 7*], the Eq. [4.3] enhanced few-parameter model fits virtually all the observed follow-on USA CoVID-19 waves, including (b) the Summer 2020 Resurgence [*Figure 11*], (c) the large Winter 2020 Resurgence [*Figures 12* and *13*], (d) the smaller Spring 2021 “*Fourth Wave*” [small *dN* / *dt* peak in *Figures 12* and *13*], and (e) the present-day Summer 2021 “*Fifth Wave*” [*Figures 14-15*].

The calculated *α*_*S*_ and *δ*_*o*_ values for this “*Fifth Wave*”, given in the *Figure 16* Table, are so small that, unabated, virtually every susceptible non-vaccinated USA person in this “minimal *Social Distancing*” and “NO Mask-Wearing” subgroup can become infected. Hopefully, as this “*Fifth Wave*” evolves, more aggressive *personal responsibility* by this susceptible sub-group will raise the pandemic-ending *α*_*S*_ and *δ*_*o*_ values.

## Data Availability

All data used is in the Public Domain or was maintained by bing.com.

## Notes

### Competing Interest Statement

The authors have declared no competing interest.

### Clinical Trial

None

### Funding Statement

No External Funding.

### Author Declarations

No Human Subjects Involved in this Data Analysis. No IRB Approval Needed.

